# A need of COVID19 vaccination for children aged <12 years: Comparative evidence from the clinical characteristics in patients during a recent Delta surge (B.1.617.2)

**DOI:** 10.1101/2021.11.05.21265712

**Authors:** Hongru Li, Haibin Lin, Xiaoping Chen, Hang Li, Hong Li, Sheng Lin, Liping Huang, Gongping Chen, Guilin Zheng, Shibiao Wang, Xiaowei Hu, Handong Huang, Haijian Tu, Xiaoqin Li, Yuejiao Ji, Wen Zhong, Qing li, Jiabin Fang, Qunying Lin, Rongguo Yu, Baosong Xie

**Author notes:** Co-first authors (Haibin Lin, Xiaoping Chen).

## Abstract

**Objective:** To evaluate the necessity of Covid-19 vaccination in children aged < 12 y by comparing the clinical characteristics in unvaccinated children aged < 12 y with vaccinated patients aged ≥ 12y during the Delta surge (B.1.617.2) in Putian, Fujian, China.

**Methods:** A total of 226 patients with SARS-Cov-2 Delta variant (B.1.167.2; confirmed by Realtime PCR positive and sequencing) were enrolled from Sep 10th to Oct 20th, 2021, including 77 unvaccinated children (aged < 12y) and 149 people aged ≥ 12y, mostly vaccinated. The transmission route was explored and the clinical data of two groups were compared; the effect factors for the time of the nucleic acid negativization (NAN) were examined by R statistical analysis.

**Results:** The Delta surge in Putian spread from children in schools to factories, mostly through family contact. Compared with those aged ≥ 12y, patients aged < 12y accounted for 34.07% of the total and showed milder fever, less cough and fatigue; they reported higher peripheral blood lymphocyte counts [1.84(1.32,2.71)×10^9/L vs. 1.31(0.94,1.85)×10^9/L; *p*<0.05), higher normal CRP rate (92.21% vs. 57.72%), lower IL-6 levels [5.28(3.31,8.13) vs. 9.10(4.37,15.14); *p*< 0.05]. Upon admission, their COVID19 antibodies (IgM and IgG) and IgG in convalescence were lower [0.13(0.00,0.09) vs. 0.12(0.03,0.41), *p*<0.05; 0.02(0.00,0.14) vs. 1.94(0.54,6.40), *p* <0.05; 5.46(2.41,9.26) vs. 73.63 (54.63,86.55), *p*<0.05, respectively], but longer NAN time (18 days vs. 16 days, *p*=0.13).

**Conclusion:** Children aged < 12y may be critical hidden spreaders, which indicates an urgent need of vaccination for this particular population.

## Introduction

Since its outbreak in early 2020, Covid-19 has caused a global pandemic, with a cumulative total of over 235,634,315 infections and a toll of tens of thousands of deaths (4,973,007 in total as of October 27, 2021). Amid the development of the Covid-19 pandemic, several mutants of the virus have been discovered, including Alpha (the B.1.1.7 in the United Kingdom), Beta (the B.1.351 in South Africa), Gamma, Lamda, Delta etc., among which the Delta variant (B.1.617.2) was first identified in Maharashtra in late 2020 [1] and spread to India and then to at least more than 180 countries or regions, making it the most aggressive widespread variant around the world. The first locally-transmitted case of Delta variant in China was reported in Guangzhou on May 21, 2021, which was introduced by imported overseas cases and resulted in a total of 167 localized outbreaks. So far, four more surges have been reported successively in Yunnan Ruili Airport, Nanjing Lukou Airport, Zhengzhou and Putian. Studies have shown that compared with the wild strain, the Delta variant shows characteristics of a shorter incubation period (4.7 vs. 6.3 days), faster transmission, higher viral load (10^6 vs. 10^4 copies) and significantly longer time for nucleic acid negativization (NAN), and deteriorates 2.98 times faster and 1.45 times more likely into a critical status for the elderly patient population aged 60 years and older [2]. The global launch of the vaccination against Covid-19 has been effective for the delta variant, with Pfizer bioNTech reducing the symptomatic population from 94% to 64% and Oxford Astrazea from 73% to 60% [3] and Moderna vaccine (mRNA 1273) preventing 81% of hospitalizations and 76% of infections and Pfizer bioNTech (BNT162B2) vaccine reducing 75% of hospitalizations and 42% of infections [4]. Several studies have also shown that vaccination provides 90% protection for frontline workers [5]. The CDC in the United States cautions that people who have not been vaccinated will face an infection risk of 11 times higher [6]. All these findings suggest that the Covid-19 nucleic acid vaccination can provide effective protection from Delta variant infection and prevent it from developing into severe diseases. On the other hand, the L452R and P681R mutants of Delta (B.1.617.2) enhance its binding to the ACEI receptor of human alveolar epithelial cells, inducing faster transmission, and trigger immune escape, resulting in compromised vaccination and impaired immunity [7-11]. Studies showed that after vaccination, the viral load in patients infected with the Delta variant was equivalent to that of unvaccinated patients [12]. Moreover, the protection against Delta variant is weaker than against other mutant strains, which may result in breakthrough cases [13], which has posed new challenges to the global efforts in containing the Covid-19 pandemic.

At present, vaccination programs for adolescents (≥12 years of age) have been approved and implemented in countries such as China, the United States, and the United Kingdom, but no mass vaccination has been conducted globally for children and adolescents under the age of 12 years due to remaining controversies over the risk-benefit issues of vaccination in children and adolescents [14, 15]. Studies have shown that children generally have favorable prognosis and only experience mild symptoms, ranging from symptoms of acute upper respiratory tract infection, mild fever, to digestive symptoms such as nausea, vomiting, abdominal pain, and diarrhea, which may account for the hesitation of a vaccination need for this particular population [16]. On the other hand, some scholars believe that the negligence of these infected children often turns them into hidden spreaders of the virus and aggravates the epidemic [17]. A study once proposed a mathematical model of SEIR disease transmission with the United Kingdom as an example and projected that if adolescents and children were included in the vaccination program, the overall COVID-19 mortality rate would be reduced by 57% and the number of long-term COVID-19 infection cases would be reduced by 75% [18]. The findings from this study suggest that vaccination in children may play an important role in reducing the overall morbidity and mortality of COVID-19. According to the current vaccination policy in China, the children under 12 years of age in this Putian surge (accounting for over 30% of the total) were not yet vaccinated against Covid-19 while most patients over 12 years of age were vaccinated, so a comparative analysis of the epidemic and its characteristics in this population will help determine the necessity of vaccination in children under 12 years of age.

### Study design and participants

The study enrolled a total of 226 individuals with positive SARS-CoV-2 Delta VOC (confirmed by SARS-Cov-2 PCR), who were admitted to the designated hospital (The Affiliated Hospital of Putian University) from September 10 to October 20, 2021. Relevant information was collected regarding the patients’ epidemiological details, clinical data, laboratory results, kinetics of viral load, and information of last negative nucleic acid test. According to the current vaccination policy of China, the individuals were divided into two groups: Group 1 (aged<12y), who had not been vaccinated, and Group 2 (aged ≥12y), most of whom had been vaccinated with Chinese inactivated vaccine from Sinopharm or Sinovac company.

### Data collection

Standardized data collection forms were employed to collect, from electronic medical records, the clinical and laboratory data. including clinical manifestations, laboratory results, treatment and prognosis. In terms of laboratory data, the cycle threshold (CT) value was measured by SARS-Cov-2 real-time PCR and the serological results included blood test, biochemical analysis, CRP, IL-6, SARS-CoV-2 IgG antibody titer, blood coagulation function, and microbe testing. The above serological tests were performed 24 hours after admission and antibody in convalescence was tested within 10 to 20 days after admission. The first day of onset was designated as the day of COVID-19 diagnosis for asymptomatic patients and the day of COVID-19 symptoms or the day of COVID-19 diagnosis (whichever was earlier) for symptomatic patients. On the 10th day of the disease course, nasopharyngeal swabs were administered every day until the negative nucleic acid testing was successively reported twice with an interval of over 24 hours. The time for NAN was defined as the duration from the first day of positive nucleic acid report to the time of the second negative nasopharyngeal swab testing of the two successive negative nucleic acid tests [19].

### Viral RNA sequencing

Fujian Provincial CDC performed the whole genome second-generation sequencing of the SARS-Cov-2 specimens by positive fluorescence real-time PCR. The epidemic strain was confirmed to be the Delta variant (B.1.617.2), with which all patients were infected.

### Clinical management

All enrolled COVID-19 patients were admitted to the hospital, quarantined, and monitored for their vital signs, body temperature and respiration. Patients whose SpO_2_ was lower than 93% were treated with oxygen therapy. Most patients were treated with Chinese traditional medicine. Some patients with fever or severe illness received the treatment of BRII 196/198 neutralizing antibody after informed consent. According to the national guidelines, glucocorticoids were only suitable for specific people, and antibiotics were used for patients with bacterial infections. In terms of disease severity, all patients were classified as asymptomatic, mild (no pneumonia by chest radiography), moderate (pneumonia by chest CT), severe (SpO_2_<93%; oxygen therapy required), or critical (RICU admission or mechanical ventilation required). Patients were discharged when the discharging standards were met [19]. The collection of clinical data was reviewed at discharge.

### Statistical Analysis

All data were analyzed with the SPSS 23 software. The data of continuous parameters were expressed as mean (variance), or median (interquartile range, IQR), and assessed by T-test for normal variables and Wilcoxon rank sum test for non-normal variables. Categorical variables were presented as frequency (percentage) and analyzed by Chi Square test and Fisher’s precision probability test. The NAN-related factors were estimated by the least-absolute method with R data analysis software. Two-tailed *p* value of less than 0.05 was considered statistically significant.

### Ethics approval

All patients signed informed consent and the ethics approval was granted by the Ethics Committee of the Affiliated Hospital of Putian University (No.202152)

## Results

### Baseline information of the patients

A total of 226 infections were enrolled, of which 77 cases were under the age of 12 years (34.07%), with 44 males (57.14%) and a median age of 9[6,9], and 149 cases were aged ≥12 years (65.93%), with 50 males (33.56%) and a median age of 39[32,50]. The children aged <12y were unvaccinated and had no underlying conditions, while 94.63% (141/149) of patients aged ≥12 years were vaccinated, of whom 9 (6.04%) received one dose of vaccine and 132 (88.59%) received two doses of vaccine and 18 (12.08%) reported coexisting conditions (11 diabetes, 6 hypertension, and 1 chronic kidney disease) (Table 1). On admission, the detected RT-PCR cycle threshold values (CT value) of children aged <12y were almost the same as that of patients aged ≥ 12y [ORF1lab value: 21.28(17.74, 26.15) vs. 23.21(17.09, 27.51), *p*=0.38; N value: 20.68(15.93, 25.33) vs. 22.50(15.59, 26.79), *p*=0.46] (Table 2).

**Table 1:** Baseline information of 226 individuals infected with SARS-CoV-2 Delta B.1.167.2.

|  | All patients<br>(n = 226) | <12<br>(n=77) | >=12<br>(n=149) |
| --- | --- | --- | --- |
| Age | 32[9,46] | 9[6,9] | 39[32,50] |
| Gender |  |  |  |
| Male | 94 | 44 (57.14% ) | 50 (33.56% ) |
| Female | 132 | 33 (42.86% ) | 99 (66.44% ) |
| Vaccinated ( % ) | 141 | 0 (0.00% ) | 141 (94.63% ) |
| One-dose vaccination | 9 | 0 (0.00%) | 9 (6.04%) |
| Two-dose vaccination | 132 | 0 (0.00%) | 132 (88.59% ) |
| Basic illness | 18 | 0 (0.00%) | 18 (12.08% ) |
| Diabetes | 11 | 0 (0.00%) | 11 (7.38%) |
| Hypertension | 6 | 0 (0.00%) | 6 (4.03%) |
| Cardiovascular diseases | 0 | 0 (0.00%) | 0 (0.00%) |
| Chronic liver diseases | 0 | 0 (0.00%) | 0 (0.00%) |
| Respiratory diseases | 0 | 0 (0.00%) | 0 (0.00%) |
| Nervous system diseases | 0 | 0 (0.00%) | 0 (0.00%) |
| Blood system diseases | 0 | 0 (0.00%) | 0 (0.00%) |
| Chronic kidney diseases | 1 | 0 (0.00%) | 1 (0.67%) |
| Metabolic disease | 0 | 0 (0.00%) | 0 (0.00%) |
| Tumor | 0 | 0 (0.00%) | 0 (0.00%) |

**Table2:**
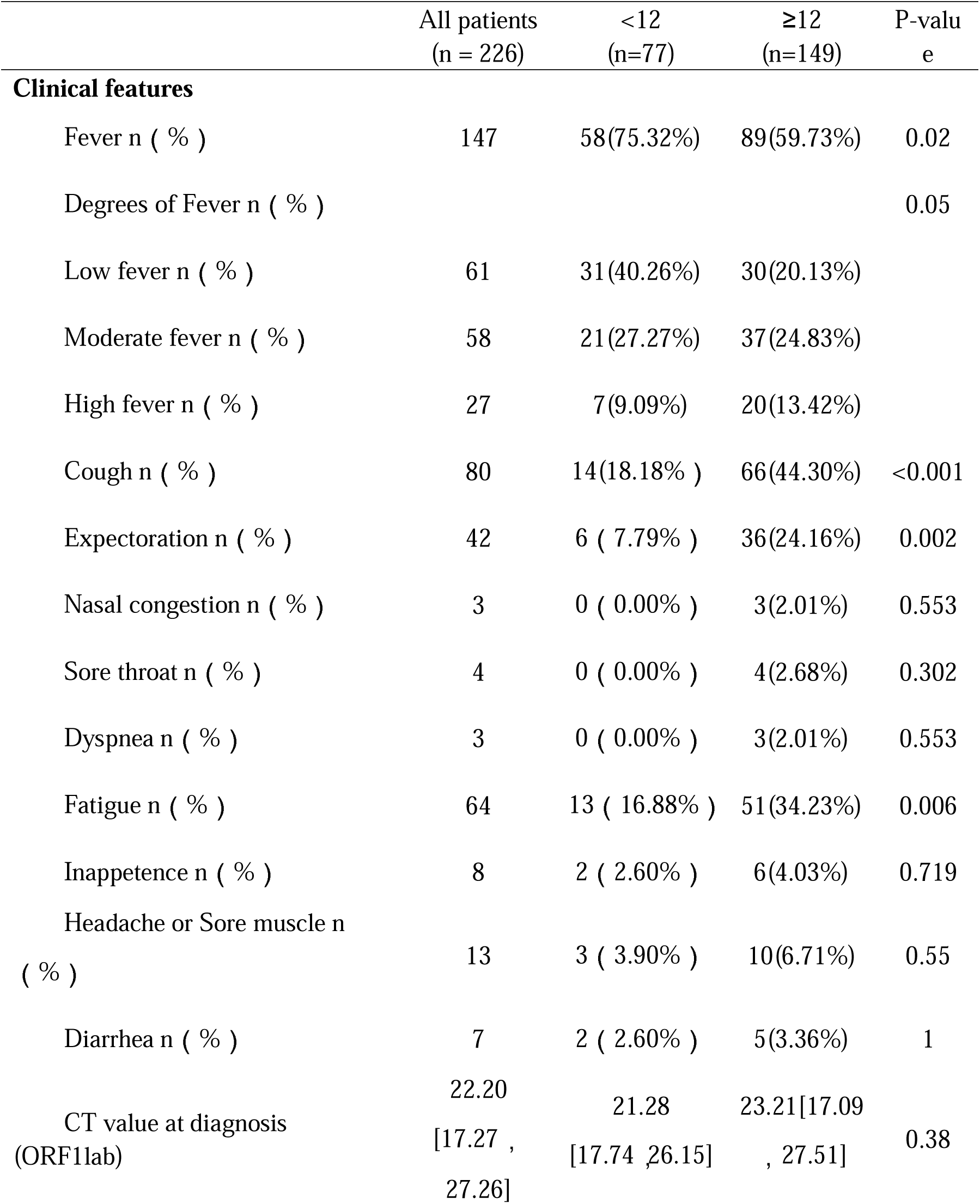
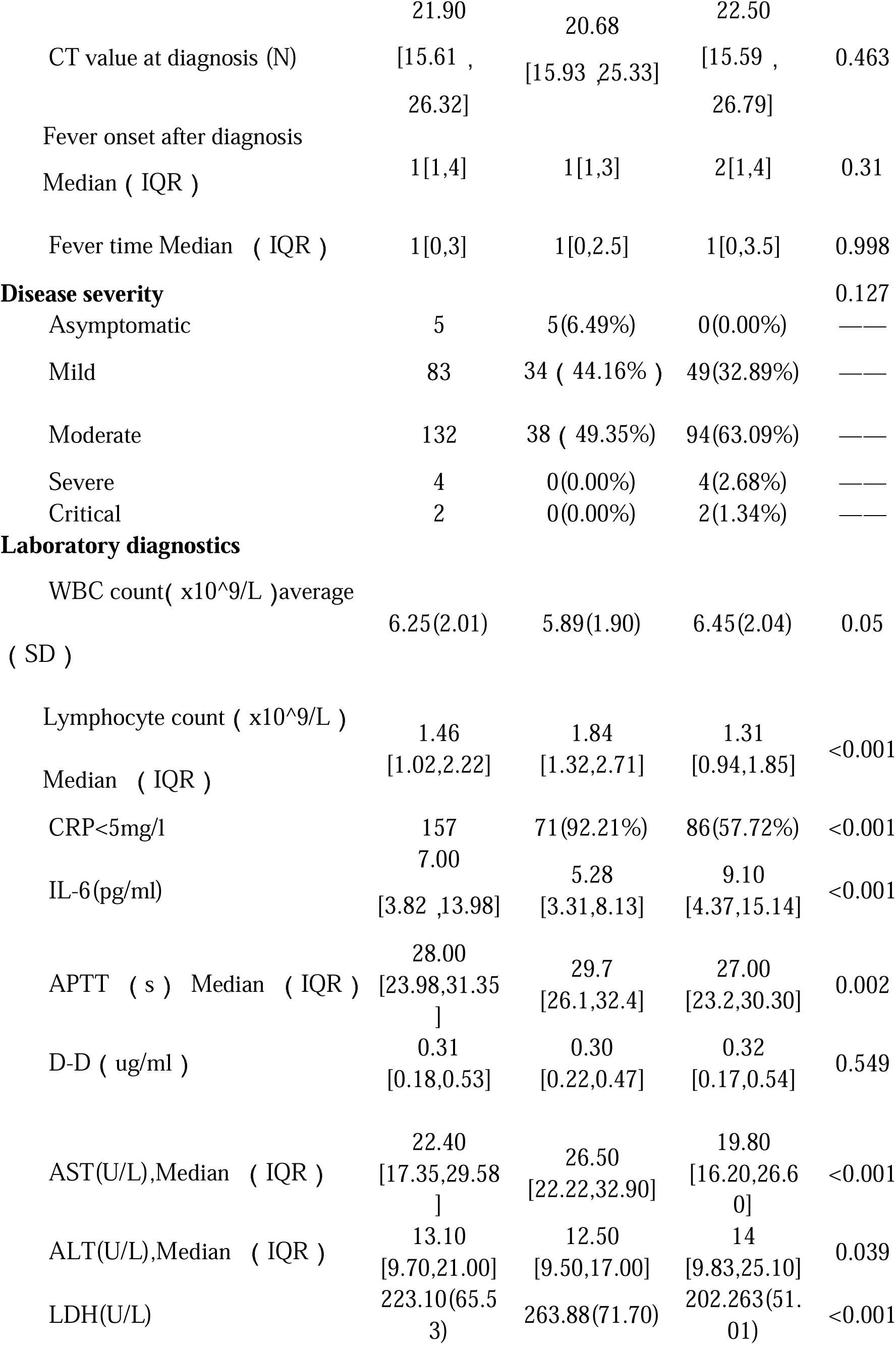
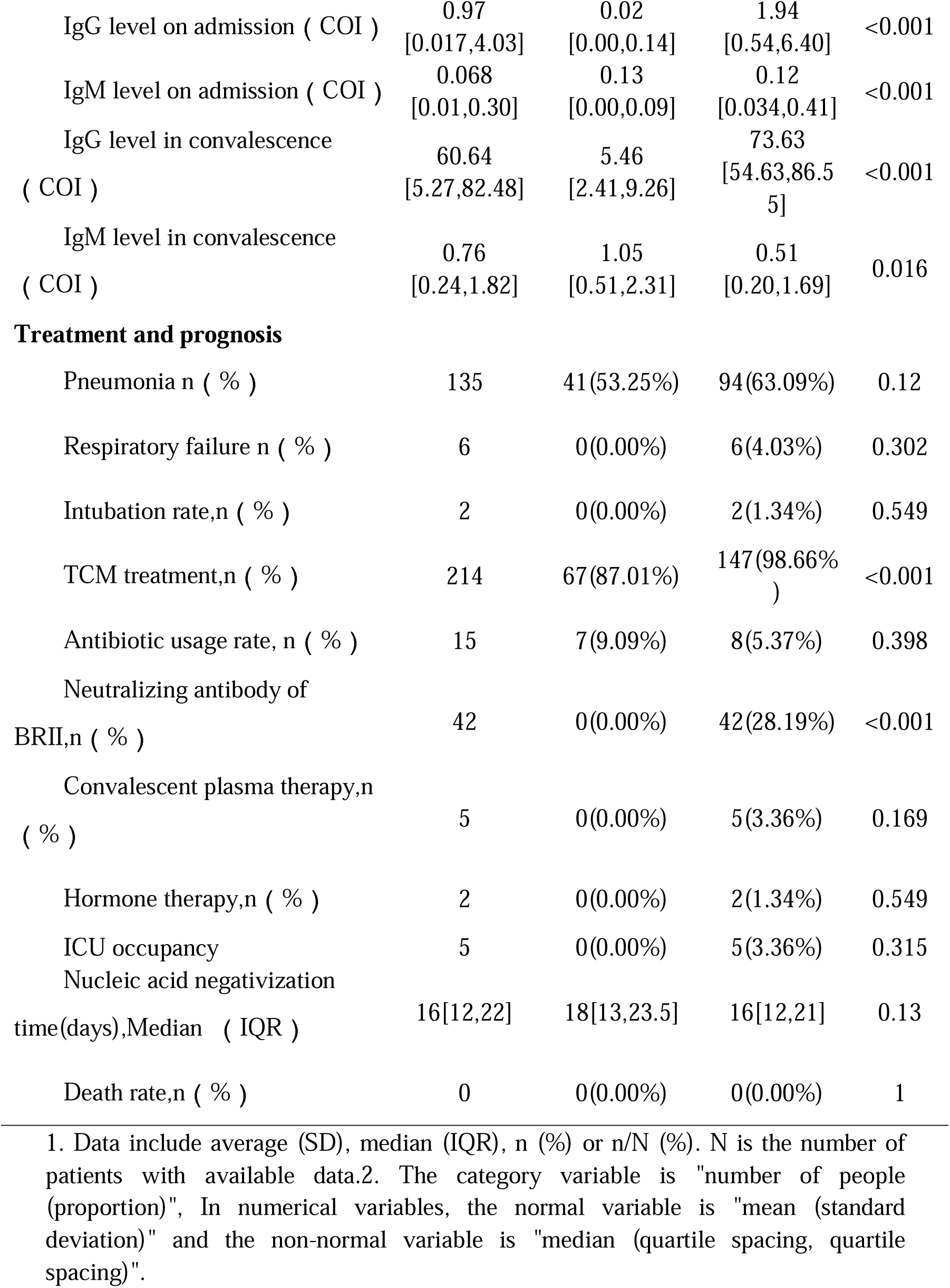
Comparative analysis of clinical features of SARS-Cov-2 Delta (B.1.167.2) infection between children aged<12y and patients aged≥ 12y.

### Transmission route of SARS-CoV-2 Delta (B.1.617.2) in Putian

In this local surge in Putian, Fujian, the first index case was a middle-aged male, who was infected during quarantine after entry into China and transmitted the SARS-CoV-2 Delta (B.1.617.2) to his two children (G1). The activities of the children in the school spread the virus to their classmates (G2), which further extended the transmission to the factories(G4) through familiy contact(G3). All of these cases were epidemiologically or genetically traced back to the first case (The specific transmission route is shown in Fig1).

**Fig1.**
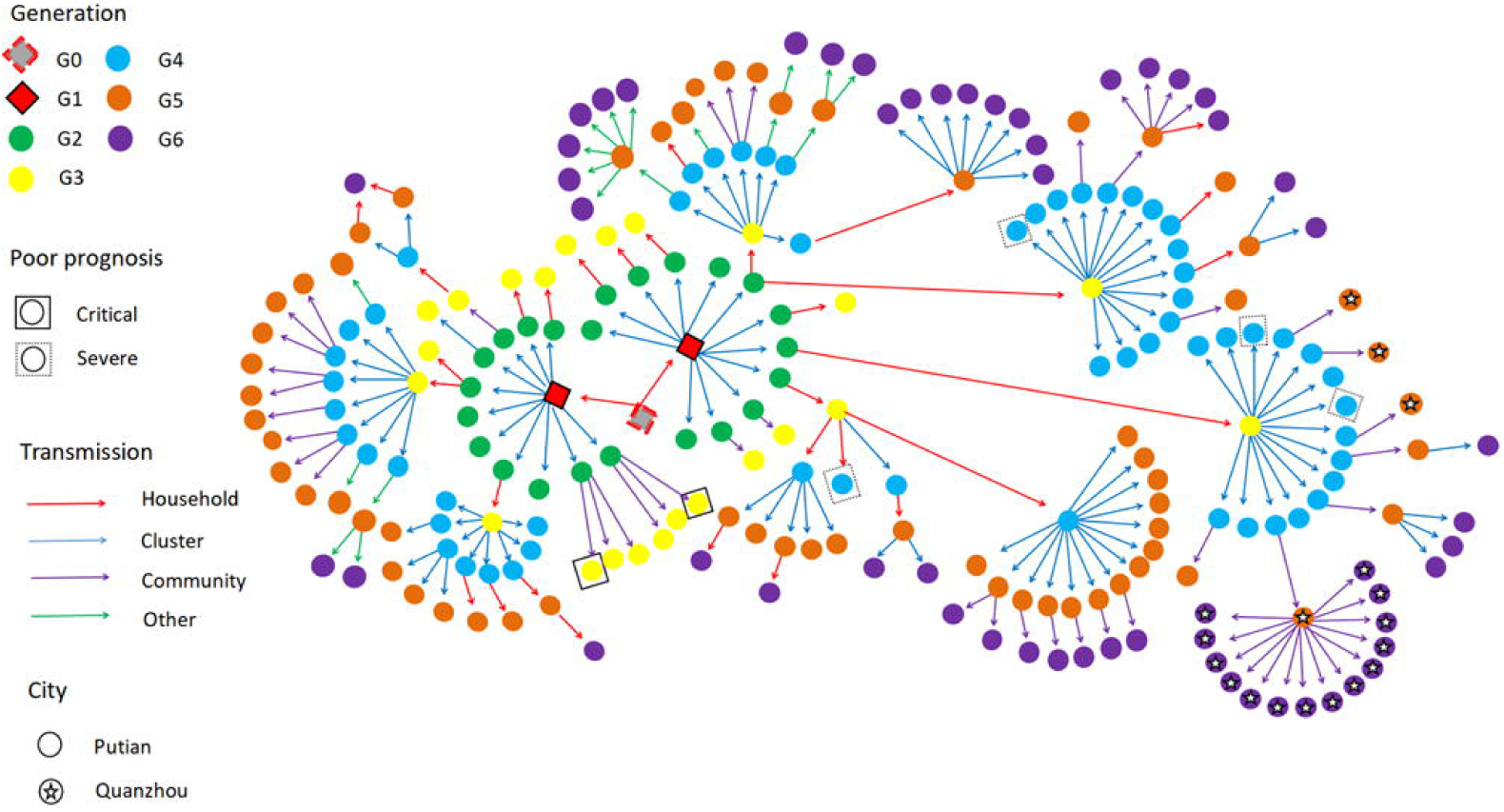
The transmission network of SARS-Cov-2 Delta (B.1.167.2) in Putian. The transmission chain of the 226 infected patients was shown in the network. Each transmission generation is shown in rhombus or circles with different colors. The first-generation patients (rhombus with black solid line, G1) were in the middle. G1 was phylogenetically linked to an imported case (rhombus with red dotted line, G0). Colored arrows indicate different transmission routes. The transmission includes household, cluster (at school and factory), community (sporadic case in the community) and others (work and social contacts). Severe (dotted line) and critical (solid line) patients were labelled with squared shapes. Asterisks indicate patients in or to other cities (For interpretation of the references to color in this figure legend, please refer to the web version of this article.)

### Comparison of clinical characteristics in patients aged <12y and those aged ≥12 y

Compared with patients aged ≥12y, 75.32% (58/77) of children aged <12y developed milder fever (*p*<0.05), minor symptoms such as cough (18.18% vs. 44.30%, *p*<0.05), expectoration (7.79% vs. 24.16%, *p*<0.05) and fatigue (16.88% vs. 34.23%, *p*<0.05). But no significant difference was found in the day of fever onset after diagnosis [1(1,3) vs. 2(1,4)] and duration of fever [1 day(0,2.5) vs. 1day(0,3.5), *p*>0.05] (Table 2).

Compared with those aged ≥12y, the younger group reported higher white blood cell count [5.89(1.90)×10^9/L vs. 6.45(2.04)×10^9/L, *p*=0.05] and lymphocyte count [1.84(1.32,2.71)×10^9/L vs. 1.31(0.94,1.85)×10^9/L, *p*<0.05), higher normal CRP rate (92.21% vs. 57.72%) (*p* < 0.05), lower IL-6 levels [5.28(3.31,8.13) vs. 9.10(4.37,15.14), *p*<0.05]; they also showed lower levels of COVID-19 antibody IgM and IgG on admission and IgG in convalescence [0.13(0.00,0.09) vs. 0.12(0.03,0.41), *p* < 0.05; 0.02(0.00,0.14) vs. 1.94(0.54,6.40), *p*<0.05; 5.46(2.41,9.26) vs. 73.63(54.63,86.55), *p*< 0.05, respectively], but higher antibody IgM in convalescence [1.05(0.51,2.31) vs. 0.51(0.20,1.69), *p*=0.016) ((Table 2,Fig2).

**Fig2.**
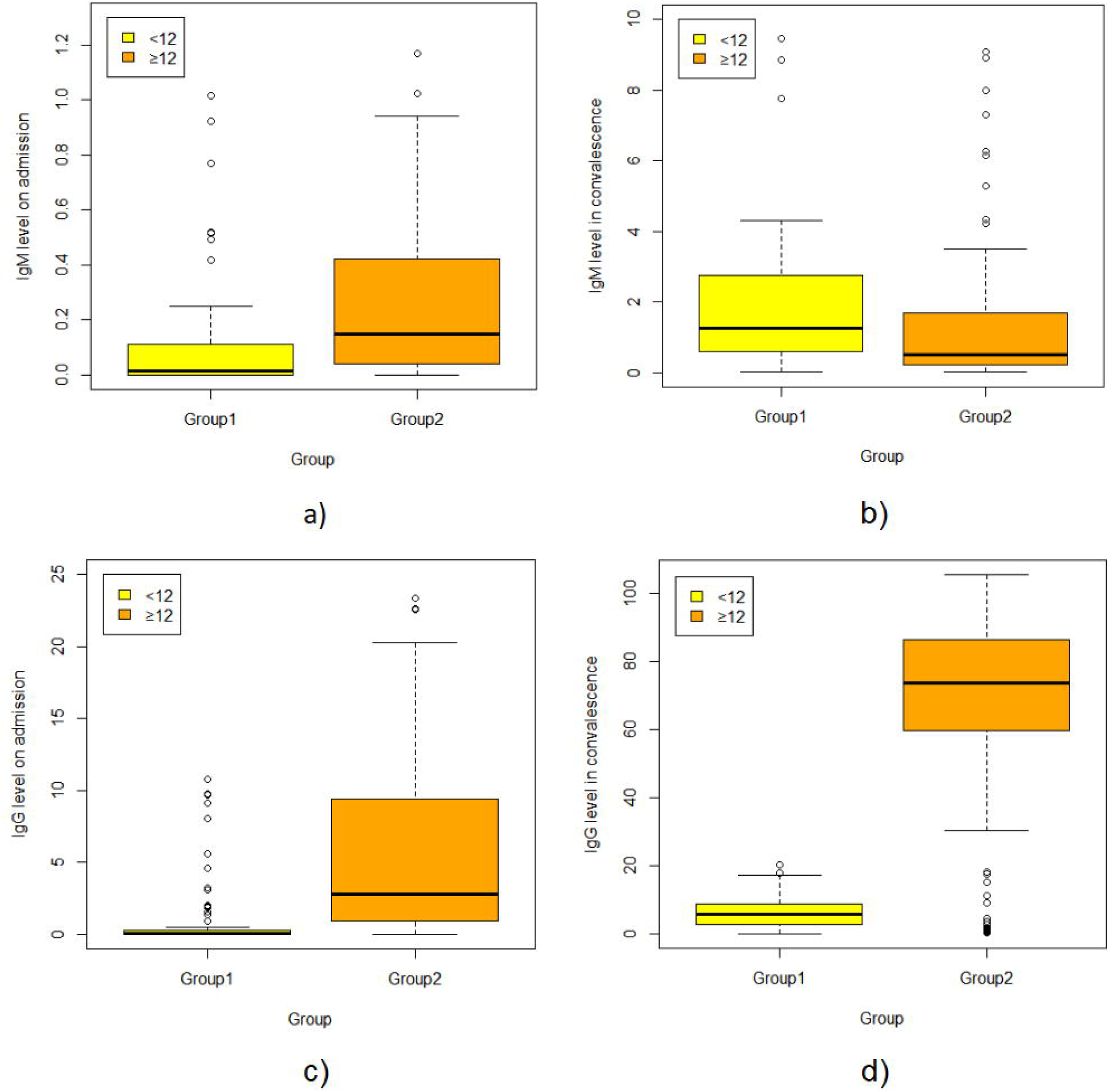
Serum SARS-CoV-2 IgM and IgG antibody levels on admission and in convalescence in the 226 individuals infected with SARS-CoV-2 Delta (B.1.167.2). Dots represent IgM/IgG level in children aged <12 years (group1) and those aged≥ 12years (group2). Box plots indicate the median and interquartile range (IQR) and the whiskers represent the maximum and minimum values. The results showed lower COVID antibody IgM and IgG level on admission, and IgG level in convalescence [0.13(0.00,0.09) vs. 0.12(0.03,0.41), *p*<0.05; 0.02(0.00,0.14) vs. 1.94(0.54,6.40), *p* <0.05; 5.46(2.41,9.26) vs. 73.63(54.63,86.55), *p*< 0.05, respectively], but higher antibody IgM level in convalescence [1.05(0.51,2.31) vs. 0.51(0.20,1.69), *p*=0.016].

Compared with patients aged ≥12y, the younger group had milder illness (*p*<0.05), with a higher asymptomatic rate (6.49% vs. 0%), more mild cases (44.16% vs. 32.89%), and fewer moderate and severe and critical cases (49.35% vs. 63.09 %; 0% vs. 4.02%, respectively); they also had a lower frequency of pneumonia (53.25% vs. 63.09%, *p*=0.05) and respiratory failure (0% vs. 4.03%) but longer NAN median [18(13,23.5) vs. 16(12,21), *p*=0.13). In the infected patients aged ≥12y, 5 patients was admitted to ICU, with a median stay length of 9 days [7, 21], 2 (1.34%) receiving endotracheal intubation; 42 (28.19%) received BRII196/198 neutralizing antibody therapy; 5 (3.36%) were given convalescent plasma; and 2 people (1.34%) received hormones treatment. However, no such treatments were administered to the children aged<12y (Table 2).

### Analysis of factors related to NAN

The data were modeled by R data analysis software. The results showed that the younger group reported a much longer NAN time when compared with patients aged ≥12y, which was negatively correlated with the IgG level on admission, the days of fever onset after diagnosis, and the nucleic acid CT value (ORF1lab) on admission, but positively with the occurrence of pneumonia, the degree of fever, and the severity of the disease (Table 3).

**Table3.**
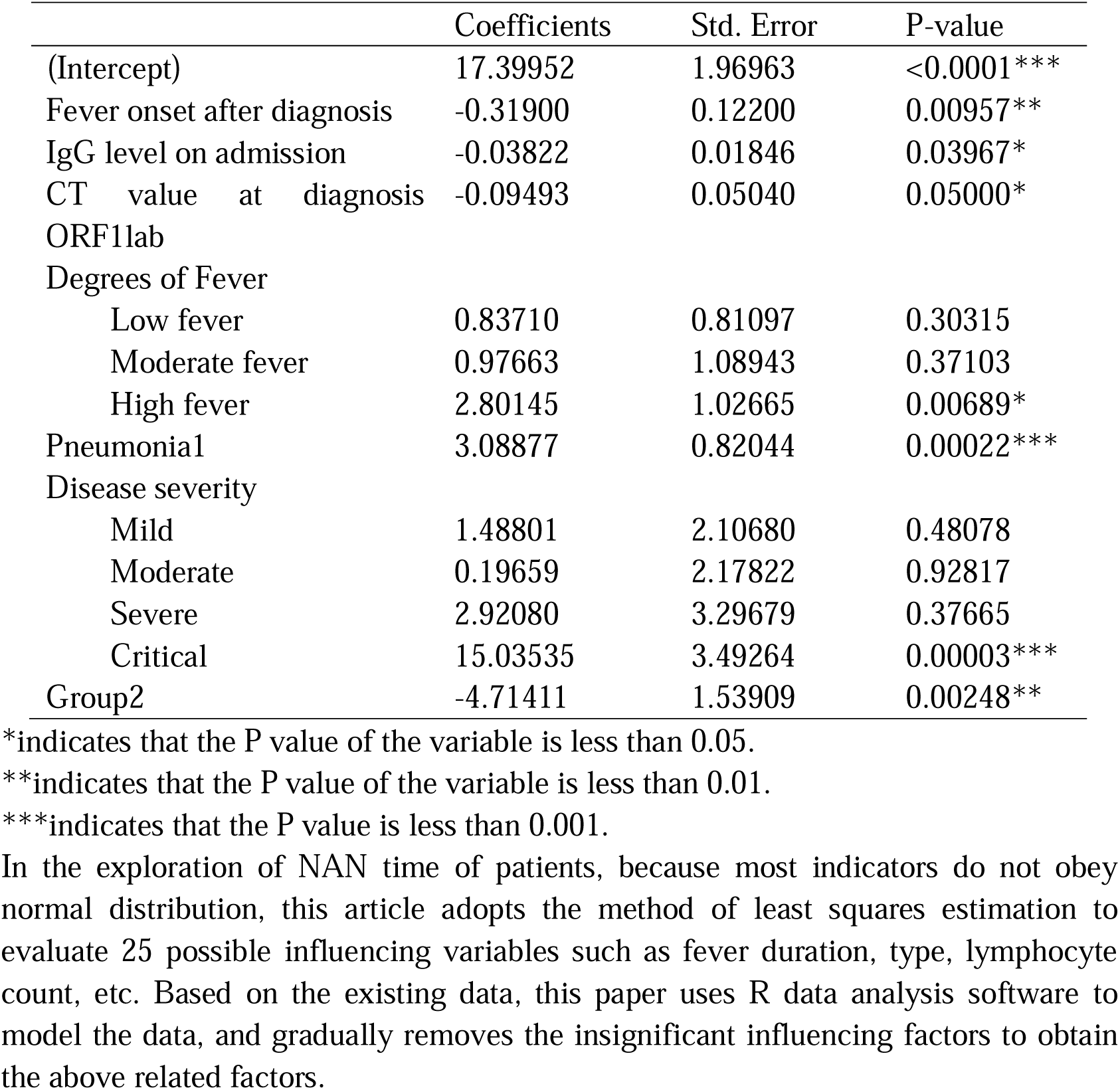
The effect factors related to NAN time for the patients with SARS-Cov-2 Delta (B.1.167.2) infection.

## Discussion

SARS-CoV-2 B.1.617.2 (Delta variant) first emerged in October 2020 and has been prevalent in the world (https://www.gisaid.org/hcov19-variants/). The transmission rate of the SARS-COV-2 Delta variant increases by 97% when compared with that of the wild-type strain [20]. Patients infected with Delta variant are twice likely to be hospitalized when compared with those with Alpha variant [21]. In the Guangdong Delta variant surge (May 2021), the viral load in patients was about 1,000 times higher than that of those infected with the original 2019 coronavirus strain [2]. The viral load and transmission speed in the Putian Delta variant surge (Sept. 10^th^, 2021) were similar to those in Guangzhou. But, in the Putan surge, children were the first and prominent spreaders, which is consistent with the findings of other studies. Previous studies show that school and family are critical transmission places for SARS-COV-2 [22, 23], in which the risk of household transmission was expected to be 60% higher for the Delta variant than for the Alpha variant [24]. In summary, children can be critical spreaders in the epidemic of Delta variant.

The Putian Delta variant surge revealed characteristics of younger individuals, milder symptoms and higher vaccination rate. Most of the patients aged ≥ 12y had been vaccinated with two doses of inactivated SARS-COV-2 vaccine while only 1/5 of the patients in Guangdong Delta surge (May 2021) were vaccinated [25]. Of note, the current study showed that for patients aged ≥ 12y, the Delta variant-related diseases were much milder in Putian than in Guangzhou surge, with fewer than one-fifth severe cases [25], which is consistent with the results of previous studies [26-28].

In addition, the median NAN time was obviously shorter than that in Guangzhou (16 vs. 19 days) [25]. Further analyses showed that the COVID-19 antibody IgG in the vaccinated population (aged ≥ 12y) increased much more rapidly and was higher than that of the unvaccinated minor group. Furthermore, a previous study documents that the nucleic acid CT value of the vaccinated group increased more rapidly than that of the unvaccinated group, indicating a much faster virus clearance by vaccination [7]. Taken together, these findings suggest that vaccination can provide sufficient protection from disease deterioration and accelerate the elimination of virus.

However, the mutation of SARS-COV-2 delta variants may cause immune escape and reduce the sensitivity to neutralizing antibodies, leading to the breakthrough infection of Delta variants even after vaccination [13]. Therefore, delta variants may still be potent for quite a long time in the future and unvaccinated children may be critical hidden spreaders due to the absence of effective immunity barrier. In our cohort, children aged <12 y were unvaccinated and accounted for more than 1/3 of the total patient population, much more than those in Guangzhou [25]. In accordance with previous studies, the symptoms of this group were mild as a whole, with milder fever, fewer other symptoms, and no severe cases, when they were compared with those who were over the age of 12 years and vaccinated [25, 29]. They had higher lymphocyte counts, as in Vono M’s study [30], which may be related to the innate immunity of this population [31-33]. Lower levels of CRP and IL-6 may indicate a lower overall inflammatory response and a good prognosis [34].

However, in contrast to the mild symptoms, the overall median NAN time of children under the age of 12 years was longer than that of those aged 12y and over. Although studies show that virus inactivation time is much shorter than the NAN time [35],the NAN time is still closely related to the elimination of virus and its infectivity. Further analysis showed that the population aged <12y and the COVID-19 IgG level on admission were two of the most important factors for NAN time. We hypothesize that as they are unvaccinated, the production of novel antibody is slower in the population aged <12y, resulting in slow virus clearance. These results suggest that if no due attention is paid to children with mild symptoms, they may carry the virus for a longer time and become hidden disseminators of Delta virus.

Previous studies have showed that a large number of RBD specific IgG memory B cells can be induced after mild infection with COVID-19 virus [36]. Children’s B lymphocytes may be activated faster and can produce much more effective antibodies [30]. The vaccination can produce more memory B lymphocytes in the younger population, which may promote antibody production and speed up virus clearance in the early stage of SARS-CoV-2 infection, so as to reduce their risk as hidden communicators. Therefore, despite the lingering medical disputes, we believe that it is of great significance to promote vaccination in the population aged <12y in order to combat the novel sars-cov-2 variant. It is worth mentioning that, presently, the clinical trials with the mRNA 1273 and bnt162b2 vaccines of Pfizer company [37], and the inactivated vaccines (coronavac) of Kexing and Sinopharm company have been carried out in children and adolescents aged 3-17y, which reported a high serum conversion rate of neutralizing antibody (96% ∼ 100%) and favorable overall safety [38, 39]. The Chinese government has recently approved the emergency use of COVID-19 inactivated vaccine in the population aged 3-17 [40]. However, as the immune system of children, especially infants, is in the process of continuous development and improvement and the relevant application data of covid-19 vaccine is limited at present, more clinical trials should be conducted to clarify the efficacy and risk. Accordingly, it is suggested that COVID-19 vaccination for children should be promoted in an orderly manner and that the effectiveness and adverse reactions of vaccination should be closely monitored [40].

Some limitations still remain in this study: 1. Although the transmission route of all cases was relatively clear and complete, data only came from local patients of Putian, so no multi-center data are accessed. 2. Although COVID-19 antibodies IgM and IgG had been detected, neutralizing antibody (RBD) was not tested and immunofunctional studies were not conducted. Further studies should be pursued to better evaluate the immune function of children age <12 in order to unravel the underlying mechanisms.

## Data Availability

All data produced in the present study are available upon reasonable request to the authors.

http:www.example.com

## Acknowledgements

The research was funded by the following grants, including Fujian Science and Technology Guidance Project: Research on the immune response system of patients infected with variant strains of the new coronavirus (2021Y0100), Fujian Natural Science Foundation (2019J01178), Central Government Guiding Local Science and Technology Development (2021L3018), Natural Science Foundation of Fujian Province (2021J01658), High-level hospital foster grants from Fujian Provincial Hospital, Fujian province, China (NO.2019HSJJ11). We thank Professor Hongzhi Huang for proofreading and polishing the manuscript.

